# Robust Estimation of Infection Fatality Rates during the Early Phase of a Pandemic

**DOI:** 10.1101/2020.04.08.20057729

**Authors:** Perikles Simon

## Abstract

During a pandemic, robust estimation of case fatality rates (CFRs) is essential to plan and control suppression and mitigation strategies. At present, estimates for the CFR of COVID-19 caused by SARS-CoV-2 infection vary considerably. Expert consensus of 0.1–1% covers in practical terms a range from normal seasonable Influenza to Spanish Influenza. In the following, I deduce a formula for an adjusted Infection Fatality Rate (IFR) to assess mortality in a period following a positive test adjusted for selection bias.

Official datasets on cases and deaths were combined with data sets on number of tests. After data curation and quality control, a total of IFR (n=819) was calculated for 21 countries for periods of up to 26 days between registration of a case and death.

Estimates for IRFs increased with length of period, but levelled off at >9days with a median for all 21 countries of 0.11 (95%-CI: 0.073–0.15). An epidemiologically derived IFR of 0.040 % (95%-CI: 0.029%– 0.055%) was determined for Iceland and was very close to the calculated IFR of 0.057% (95%-CI: 0.042– 0.078), but 2.7–6-fold lower than CFRs. IFRs, but not CFRs, were positively associated with increased proportions of elderly in age-cohorts (n=21, spearman’s ρ=.73, p =.02).

Real-time data on molecular and serological testing may further displace classical diagnosis of disease and its related death. I will critically discuss, why, how and under which conditions the IFR, provides a more solid early estimate of the global burden of a pandemic than the CFR.

## Introduction

In the early phase of a pandemic caused by a novel pathogen, it is difficult to estimate the final burden of disease. In the case of the ongoing pandemic caused by SARS-CoV-2, it has been proposed, on the one hand, that it will be the most serious seen for a respiratory virus since the 1918-1919 H1N1 Influenza pandemic^1^. This pandemic contributed to the premature death of 1 percent of the world population at the time being^2^. On the other hand, despite all non-refutable morbidity, COVID-19 may fall short of provoking a comparable impact on mortality as the seasonal Influenza, which is estimated to contribute to 389,000 deaths per year on average^3^. Experts conclude that there is still a range for CFR of 0.1 up to 1.0, which practically spoken is reflecting the margin from normal seasonal Influenza to the lower boundary CFR estimate of 1918-1919 H1N1 Influenza^4^.

Case Fatality Rates (CFR) can be helpful to critically control and reflect the outcome of robust modelling to estimate the global burden by mortality^1^. During the phase of an outbreak CFRs are preliminary and should be communicated and used with caution^5^. Even in the case of the well-known and frequently studied seasonal Influenza A, it is a matter of debate on how to estimate a CFR during the phase of the pandemic, which could be calculated by the total number of “deaths” divided by the number of “cases”. There are ongoing discussions, what can and should be regarded as a “case” and a reasonably causal-related death^6^. A “case” could ideally be a confirmed case of the infectious disease according to strict diagnostic guidelines, requiring symptoms and confirmatory testing. Moreover, the death would ideally be a causally related death and not a death caused by superinfection over the course of hospitalization, for instance.

Whether we impose less or more strict guidelines to define a suitable “case” and its causally related death, either way will inevitably introduce selection bias. This may lead to both, either substantially higher, or substantially lower estimation of CFRs^6,7^. On the one hand, it can be argued that a strict procedure for confirmation of official cases and deaths may underestimate the effect of disease on mortality, since we will miss out both, deaths and cases, for a significant proportion of the population ^7^. On the other hand, an infection with a pathogen of mild up to medium virulence, like SARS-CoV-2, can be completely asymptomatic, or may cause only minor symptoms to a majority of infected persons^8,9^. At present the size of the denominator of total community infections is unknown^4^, but at least there is increasing evidence that a major part of the younger population, rarely become symptomatic^10^ and even more rarely could die of the disease, if diagnosed with COVID-19^9^.

At the beginning of a pandemic with a novel pathogen central aspects of previously acquired immunity or genetic resistance against viruses^11^, or other pathogens are often unknown or only vaguely explored. If they existed, but were not taken into account, the estimated CFR would not reflect the burden of an infectious disease on a macro perspective. In such a setting looking preferentially at those, who have the full-symptomatic disease or are subjected to the surveillance system, can severely overestimate the burden of disease. Therefore, in the early phase of an outbreak, infectious disease epidemiologists will rather base their estimates of the potential burden of a pandemic rather models requiring basic assumptions on the basic reproduction number R_0_, the latent period of the infectious period, and the interval of half-maximum infectiousness and many other factors. These all need to be derived from early field studies that often need to be conducted under sub-optimal conditions, in the heat of a pandemic^10^. These basic assumptions of epidemiological key figures are then used for modelling the almost uncharted^1,11,4^. From a stochastic point of view, such modelling is prone to an exponential propagation of imputation error, which can principally be controlled for and reported with these models^12^. However, as in the present case such error propagation will finally arrive at conclusions via modelling, that are indeed based on assuming merely all or nothing^4^ and thus indeed would need to be communicated with uttermost caution to the public and foremost health politicians^5^.

Under these circumstances, it could be helpful to look at alternative ways to asses a robust figure for CFR as a typical, hard to predict estimate for global burden of a pandemic. Alternative ways may allow interdisciplinary abductive reasoning to arrive at an estimate for the global burden of a disease. Particularly under non well-defined and dynamic circumstances, abductive reasoning can be more helpful than the best medical evidence employing inductive statistical inference, alone^13^.

There has just recently been a promising approach to arrive at a more robust measure of mortality^14^. This involved calculating an Infection Fatality Rate (IFR), which takes not only the asymptomatic population and their relevance for mortality into account, but also adjusted for censoring and ascertainment bias. However, this approach required again an immense workload on retrieving and curating valid data retrieved, from cohorts studied under specific conditions, and making pre-assumptions, which is again an approach prone to error propagation.

Here I will deduce that an IFR adjusted for selection bias in favour of more morbid persons, can be determined with the help of available official data in conjunction with the testing figures. I will show that the IFR adjusts the CFR for some essential sorts of bias. In order to cross validate the computed figures, I will estimate infection fatality from an ongoing large-scale testing pool of citizens representative for the general population in Iceland. At the time of finishing the dataset (4^th^ of April) 3.7 % of the general population in the representative cohort and 2.9% of the typically symptomatic part of the population had been tested^15^. Finally, I will compare how CFRs and the calculated IFRs are suited to reflect essential epidemiological aspects already known to be associated with COVID-19.

## Methods

The estimation of an IFR is based on two different and - regarding the influence of selection bias - divergent procedures to calculate a CFR from infection-related population data. The first formula (1) is a variation of the non-adjusted CFR in the following termed “classic CFR”, which divides the sum of deaths by the sum of cases on a given day. Formula (1) takes into account the persons that passed away or recovered, and days from reported positive testing (d_0_) to death (d_n_) as period (d_0-n_) into account.

For a given time interval of n days from (d_0-n_) at d_0_ sums of test positives (TP_0_), deceased persons (DP_0_) and recovered persons (RP_0_) are given and at d_n_ sum of deceased persons (DP_n_) are known. Then a case fatality ratio CFR for the interval d_0-n_ as CFR_n-0_ can be calculated:

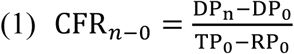

For the data provided by Johns Hopkins University (JU) the recovered persons (RP_0_) can be included, while for the data provided by the European Center of Disease Control (ECDC) a more simplified version can be calculated substituting TP_0_ − RP_0_ with TP_0_. Therefore, data of the ECDC were mostly used for quality control aspects. They served to critically revising some negative case and death numbers in the data set of JU for numbers that had been officially reported at one day to the WHO, and the ECDC, but then were then corrected later on by national health authorities. This was for instance the case for the data of Iceland.

As I mentioned in the introduction the denominator of total community infections is unknown, but a rather critical factor of uncertainty under the given ongoing pandemic. Here I will put this into a likewise simple mathematical term by calculating the CFR as a CFR’, taking this unknown denominator into account. For the beginning of a pandemic RP_0_ is often low or zero. If the total number of tests (N_t_) conducted at d_0_ is known (N_t_) an CFR’ can be calculated, by taking the total population N_p_ of the country or region into account, in which testing had been conducted. Noteworthy, this is not a typical formula to calculate a CFR, since it may tend to underestimate the true fatality until the infectious disease has stopped spreading in the population. However, just like formula 1, at the end it depends only on registered death of persons and number of cases and it is subject to the same factors that add bias on the estimation of a mortality figure, but only in the opposite direction. Moreover, since the size of the denominator of total community infections is unknown but appears to be highly relevant, this equation now puts the CFR into a context with the general population. The prevalence (p) for non-recovered infected persons in a total population (N_p_) can be calculated:

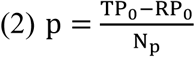

Based on the prevalence of infected persons in a population a CFR’ for the case fatality for the interval d_0-n_ (CFR’_n-0_) can be calculated:

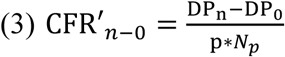

Both formulas (1) and (3) will have their shortcomings. It will be briefly discussed why (1) will most likely overestimate and (3) underestimate an IFR under the unavoidable premise that official testing will tend to test more morbid persons. In the equations (1) and (3) the pool of newly infected persons is subject to selection bias. Formula (3) typically underestimates IFR, because the prevalence p of active cases is typically determined too high to be generalized to the total population. During an outbreak, this is unavoidable for testing strategies solely based on the health care systems, since guidelines for testing require - or at least favour - preferential testing of persons with an accumulation of risk factors, like specific disease-related symptoms, or stay or visit at an endemic region. Accordingly, active cases are overrepresented in a relatively too small pool N_t_ to be representative for the p in the general population N_p_. Furthermore, some countries or regions may have limited resources when a pandemic proceeds and will adjust their testing guidelines to detect positive cases with as few N_t_ as possible. To control for this distortion by selection bias aiming at enriching for positive cases in the test pool N_t_ we need to adjust the overestimated prevalence p with the unknown factor *f* to turn the CFR′_*n*−0_ into the IFR_n-0_

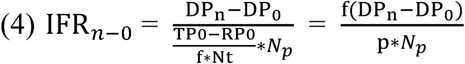

In (1) the calculated CFR is determined rather too high and does not represent an IFR, because of just the same distortion factor *f* as in (3). Since testers selectively address the pool of diseased persons, into the pool of persons tested N_t_, they will therefore also increase the risk of death that would be representative for the population of all infected persons, which should be reflected by our IFR. Likewise, we will need to correct CFR_n-0_ by the same factor *f* as in (3). In this case, IFR_n-0_ can be calculated as:

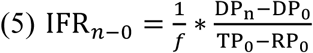

At this point, I suggest the reader to jump to Table 1 in the results section, to understand better, why CFR is divided by *f* and CFR’ multiplied with *f*. Typical potential distortion factors, like the gross domestic product of a country, but first and foremost the age composition of the population, are inversely correlated with CFR or CFR’. To adjust these two equations for all the factors that act in divergent ways on the figures calculated by (1) or (3), we can now solve the equation below for the common distortion factor f to adjust the CFRs calculated in (1) and (3) and estimate an IFR_*n*−0_. Solving for f:

**Table 1:** Spearman’s ρ (n=89 days for 10 countries) and p-Values for associations of CFR and CFR’ with age cohort distribution, gross domestic product (GDP), test number growth and test positive ratio.

| Type | against | $\rho$ | p-Value | Type | against | $\rho$ | p-Value |
| --- | --- | --- | --- | --- | --- | --- | --- |
| CFR | Male (25+) | 0.153 | 0.1025 | CFR' | Male (25+) | -0.3489 | 0.0008* |
| CFR | Male (50+) | 0.3409 | 0.0002* | CFR' | Male (50+) | -0.4986 | <.0001* |
| CFR | Male (75+) | 0.4121 | <.0001* | CFR' | Male (75+) | -0.2874 | 0.0063* |
| CFR | Male (80+) | 0.4121 | <.0001* | CFR' | Male (80+) | -0.2874 | 0.0063* |
| CFR | Female (25+) | 0.0723 | 0.4427 | CFR' | Female (25+) | -0.7506 | <.0001* |
| CFR | Female (50+) | 0.2116 | 0.0232* | CFR' | Female (50+) | -0.6238 | <.0001* |
| CFR | Female (75+) | 0.4274 | <.0001* | CFR' | Female (75+) | -0.4607 | <.0001* |
| CFR | Female (80+) | 0.4442 | <.0001* | CFR' | Female (80+) | -0.4549 | <.0001* |
| CFR | GDP | 0.1466 | 0.1181 | CFR' | GDP | -0.6194 | <.0001* |
| CFR | GDP/Person | -0.1673 | 0.0739 | CFR' | GDP/Person | 0.1337 | 0.2116 |
| CFR | Test no. growth | 0.2652 | 0.0077* | CFR' | Test no. growth | 0.5358 | <.0001* |
| CFR | Tests positive % | 0.2277 | 0.0319* | CFR' | Tests positive % | 0.0112 | 0.9279 |

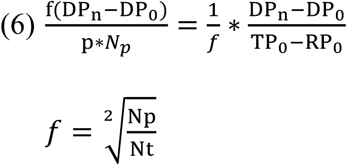

Accordingly, CFR adjusted with the factor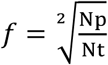, resembles an IFR, which can be calculated by equations 5 or 4, with both equations delivering the same outcome. To compare estimates of the IFR I analysed the data on cases, deaths and recoveries published in real time^16^ and once daily after correction by JH^17^. Data between the 5^th^ and 22^nd^ of March were combined and validated with the respective data from the ECDC^18^. For the numbers of death I used the data for the training data set in its final corrected version from JH for the end of the 29^th^ of March, for the enlarged final dataset the 31^st^ of March, and for the data set for the validation with the epidemiolocal project in Iceland the version 4^th^ of April^17^.

For the calculation of the epidemiologically derived IFR_deCode_ we did not need to apply the correction factor *f*, because the prevalence p in the population for revealing a positive test has been determined experimentally as p_deCode_ which was used to determine the IFR for the general population of Iceland with the formula:

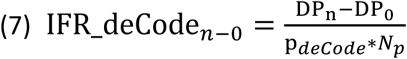

This formula is not relying anymore on cases reported in the official databases of JH or ECDC and it served as a cross-validation figure for the IFR and the CFRs, which are solely based on these data and the population data of Iceland in the validation part of the results section.

For the final analysis in the training and the final dataset only countries were included which reported at least one death by the 22^nd^ of March in the ECDC and the JH database. To avoid undue variance by too few case numbers only data points from countries were included with more than a sum of 50 cases, and at least two official reports of the numbers of tests performed in the period 5^th^ of March till 22^nd^ of March. Data on test frequencies were originally obtained from the open-source “Our World in Data”^19^. Population data of nations were imported from World Bank^20^ and data on gross domestic products and age cohort compositions from the UN^21^ in their version for 2018 to be comparable with the population data from 2018 or as age cohort estimates for 2020. For 14 countries, sufficient data were controlled, corrected and updated with information from the official data source pages of the national health agencies as listed in Supplemental Table 1.

Two more datasets were obtained for a validation study. First, data from a testing cohort of the normal population of Iceland led by the genetic company deCODE^15^. The project is planned as a clinical project with ethical permit and so far, there is to the best of my knowledge no other data available on representative study cohorts from populations without being pre-selected for pathological symptoms. Second, data mining for a final enlarged dataset was done on the pages of the official national health agencies, Wikipedia, and within the data mining community on GitHub using archived webpages if necessary in order to enable a large-scale cross-country assessment and comparison of IFR-values. In subset-analysis on multiple entries for the log-normalized cumulative testing data, there was a Pearson’s correlation coefficient between data entries across the three different data sources > 0.99 (data not shown).

Most of the data nested into groups showed signs of unequal variance by Barlett and Levene testing. Therefore, they were log-normalized and in case data included 0 an offset of 0.01 was added before log-normalization, taking care that this did not distort the distribution of data, as analysed by Shapiro-Wilk-testing. Normal distributed data with equal variance across groups were then compared using one-way ANOVA *F*-testing. Global significance was followed up by all-pairs Tukey-Kramer Testing as post-hoc test. For reporting, data were de-normalized adding the offset where necessary and reported as means with 95-% CIs, if not specified otherwise. If signs of non-normal distribution or unequal variance prevailed, a Wilcoxon-Test on rank sums for group comparisons, or a Spearman’s rank correlation coefficient ρ was reported. For the testing data set descriptive median values and their interquartile – ranges were reported.

## Results

### Implications from the Training Dataset

The combined datasets from ECDC and JH contained information on deaths, cases, and in the JH dataset information on recoveries. For ECDC 4,500 data entries from 179 countries starting on the 31^st^ of December 12.2020 and for JH 3,159 entries from the same countries starting on 21^st^ of January 2019 were combined. Cumulative cases and deaths were significantly correlated between both data sets (Pearson’s r > 0.99, p < 0.0001, for both, data not shown). No data on test frequencies were reported in the official international data repositories. From the platform “Our World in Data” 77 different data entries for 49 countries were retrieved. After combination with the official data on test figures, fourteen countries fulfilled inclusion criteria. Testing data for these countries were controlled by visiting the official test report pages of the fourteen countries (supplemental Table 1), which enabled adding another 77 data points.

The calculation of the classic CFR by dividing deaths by cases revealed a large range for the respective medians of the 14 countries and CFR calculated for the period from the 22^nd^ to the 29^th^ of March surpassed 100% for 9 countries (Fig. 1). Excess mortality was present throughout most points in time for Italy, UK, France, the Philippines, and Canada, except for one data point, which was related to a period from case to death of only one week. Excess mortality or mortality too close to the point, or even with the point of death is a bias, which will not be corrected by the factor *f* and will inevitably lead to an overestimation of CFR with both formulas (1) and (3).

**Fig.1:**
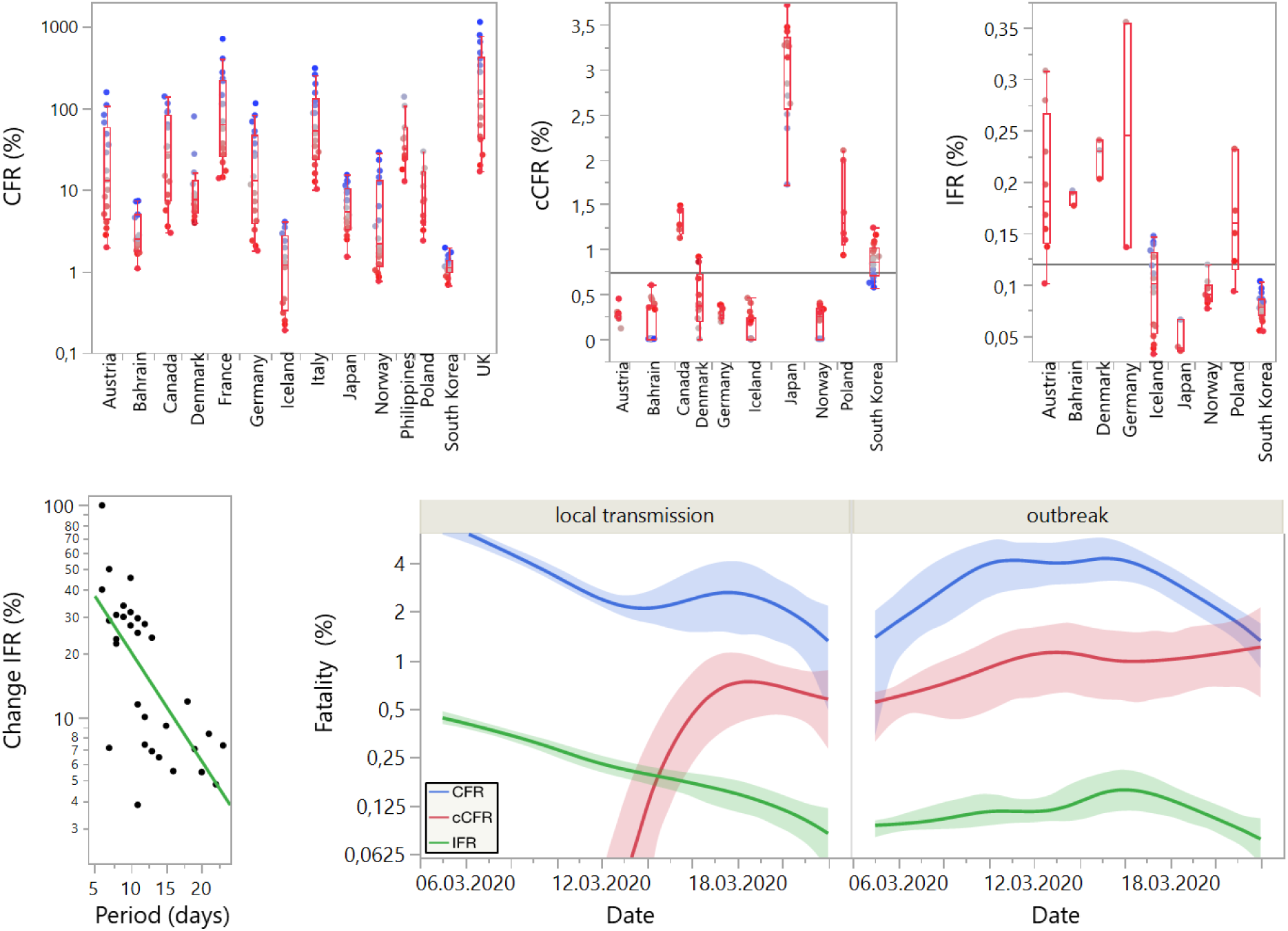
Upper left graph shows CFR values for 14 countries for a time-span of 7 – 24 days. Colour merges from **blue** (5^th^) over **grey** 13^th^ to **red** (22^nd^ of March) for the day of the cases (TP_0_) indicating increase in period (days) up to a maximum of 14 days with the 29^th^ of March as end point for death (DP_n_). Values > 10% were excluded from further analysis as explained in the results section. Upper right shows the classic CFR calculated as total deaths at one day divided by active cases at same day for the remaining 10 countries. The classic CFR is the higher, the more recent data were assessed (legend for all upper parts). IFR was calculated for the 10 remaining countries and is shown at lower left to vary also depending on period. While the IFR increases with period, this increase declines significantly with increasing period as shown in the lower right. In comparison with the CFR (blues curve) with 95% CI on splined data with moderate λ, the classic CFR red curve, the IFR green curve shows higher values in countries that have the WHO status (25.03.2020) “in local transmission”. Values reached those of the countries either the status “outbreak” after the occurrence of the first reported death.

CFR values above 100% are theoretically impossible, while CFR values over 10% are at least highly unrealistic. Therefore, I excluded these data points with a CFR > 10 from further analysis. Noteworthy, this led to the removal of all data points from Italy (classic CFR = 6.99,) France (2.02), UK (1.79) and Philippines (7.51). Noteworthy, selection did not exclude Japan with a classic CFR of 3.14 (Table 2).

**Table 2.** shows the medians and quartiles of the three different CFR values and the IFR

| Country | Classic CFR (%) and median quartiles |  |  | CFR (%) median and quartiles |  |  | CFR' (%) median and quartiles |  |  | IFR (%) median and quartiles |  |  |
| --- | --- | --- | --- | --- | --- | --- | --- | --- | --- | --- | --- | --- |
| Austria | 0.27 | 0.23 | 0.30 | 4.50 | 2.93 | 7.71 | 0.01 | 0.01 | 0.01 | 0.18 | 0.14 | 0.27 |
| Bahrain | 0.00 | 0.00 | 0.37 | 2.50 | 1.76 | 4.94 | 0.02 | 0.01 | 0.02 | 0.19 | 0.18 | 0.19 |
| Denmark | 0.37 | 0.20 | 0.72 | 6.37 | 4.58 | 8.13 | 0.01 | 0.01 | 0.01 | 0.23 | 0.20 | 0.24 |
| Germany | 0.27 | 0.23 | 0.37 | 3.67 | 2.12 | 6.76 | 0.01 | 0.01 | 0.01 | 0.25 | 0.14 | 0.36 |
| Iceland | 0.00 | 0.00 | 0.24 | 1.23 | 0.33 | 2.76 | 0.01 | 0.01 | 0.01 | 0.10 | 0.05 | 0.13 |
| Japan | 3.14 | 2.56 | 3.36 | 3.96 | 2.98 | 6.65 | 0.00 | 0.00 | 0.00 | 0.04 | 0.04 | 0.07 |
| Norway | 0.25 | 0.00 | 0.35 | 1.65 | 0.99 | 3.04 | 0.01 | 0.01 | 0.01 | 0.09 | 0.08 | 0.10 |
| Poland | 1.29 | 1.06 | 2.02 | 4.41 | 2.98 | 7.25 | 0.01 | 0.00 | 0.01 | 0.16 | 0.12 | 0.23 |
| S. Korea | 0.86 | 0.70 | 1.01 | 1.11 | 0.96 | 1.37 | 0.01 | 0.01 | 0.01 | 0.08 | 0.07 | 0.09 |

Though it is plausible that conducting more tests per day, can contribute to artificially increasing both types of CFRs, countries may also respond with increasing their test numbers, once they notice increases in the test positive ratio as a sign for focusing testing too much on the more morbid part of the population (Table 1, last line).

In Table 1 the data points with CFRs > 10 % had been excluded, already. Compared with CFR < 10%, the excluded CFRs > 10% showed with a median of 10.9% a 3.3-fold higher test positive ratio (chi^2^ = 17.6, df = 1, p< .0001) and with a median of 0.007% on average 4.0-fold fewer tests per inhabitant (chi^2^= 30.7, df = 1, p = .0001).

Spearman’s ρ and p-Values for the correlation of CFR and CFR’ with age cohorts and gross domestic product (GDP) and test growth ratios are shown in Table 1. The test growth ratio reflects the increase in tests - not cases - per day and was positively associated with CFR (Spearman’s ρ = 0.27; p = .0001) and CFR’ (Spearman’s ρ = 0.54; p = .0001). All other values were inversely associated with both CFR-Types in line with the hypothesis (see Methods).

In the following we will use *f* as an adjustment factor for determining the IFR_*n*−0_ of SARS-Cov-2 and comparing the obtained values with the three different CFRs (Tab. 2). The median IFR values of the nine remaining countries lie in a close margin between 0.07 for South Korea and 0.20 for Denmark, while classic CFR and CFR values still show high variance for the remaining countries. Especially for South Korea and Japan as two comparable countries that are in a phase of stagnation the CFR values calculated are roughly 3-5 times higher and still rising and they are without correction by *f* not in the margin of the current expert consensus for a CFR for COVID-19 (Table 2).

In contrast to the classic CFR, CFR and CFR’ the IFR values, including Japan, were in the lower range of expert consensus. As the classic CFRs the IFR can depend on the length of the period from cases included to the deaths they are related to Figure 1 (lower part). A correlation of the percent increases from day to day with increasing periods in days for all IFRs computed for the 9 countries shown in Table 2, showed a significant negative trend (Pearson’s r = 0.69; n = 31; p = .0001). Therefore, dependence on the period between cases and deaths seemed to become more moderate over time and was rather related to the state of the pandemic categorized as “in local transmission” or “outbreak” (Fig.1, lower right). As can be observed from the curve of the classic CFR (cCFR), data, which rely on cases assessed before the first death related to the outbreak was registered, tended to be either far too low (cCFR), or to high CFR and IFR. Therefore, for the following validation of data from Iceland and the evaluation of the final validation dataset, care was taken to include data after the first death reported which ideally also reflected at least a period of 9 days for calculation of the IFRs.

### Validation of the IFR with testing data retrieved from the general population in Iceland

For validation of my procedure, I analyzed data from two different testing cohorts in Iceland^14^. Up to the last data entry for the 4^th^ of April, 3.7 % of the general population in the representative cohort had been tested for SARS-CoV-2 by the genetics company deCODE (deCODE). Additionally, a second, rather typical test cohort of persons with increased risk of infection, representing 2.9% of the total population, had been tested by the Directorate of Health in Iceland via the laboratory of the National University Hospital Iceland (NUHI). Only data were included following the first death on the 17^th^ of March and allowing for at least a 9 day period of the IFR.

Prevalence of SARS-CoV-2 positive tested persons was 13.0 fold-lower (CI: 12.0-14.0) in the deCODE collective, which was highly significantly different from the correction factor *f* at 6.07 (CI: 5.63 - 6.56, *F* = 222, df = 1, p = .00001; Figure 2).

**Figure 2:**
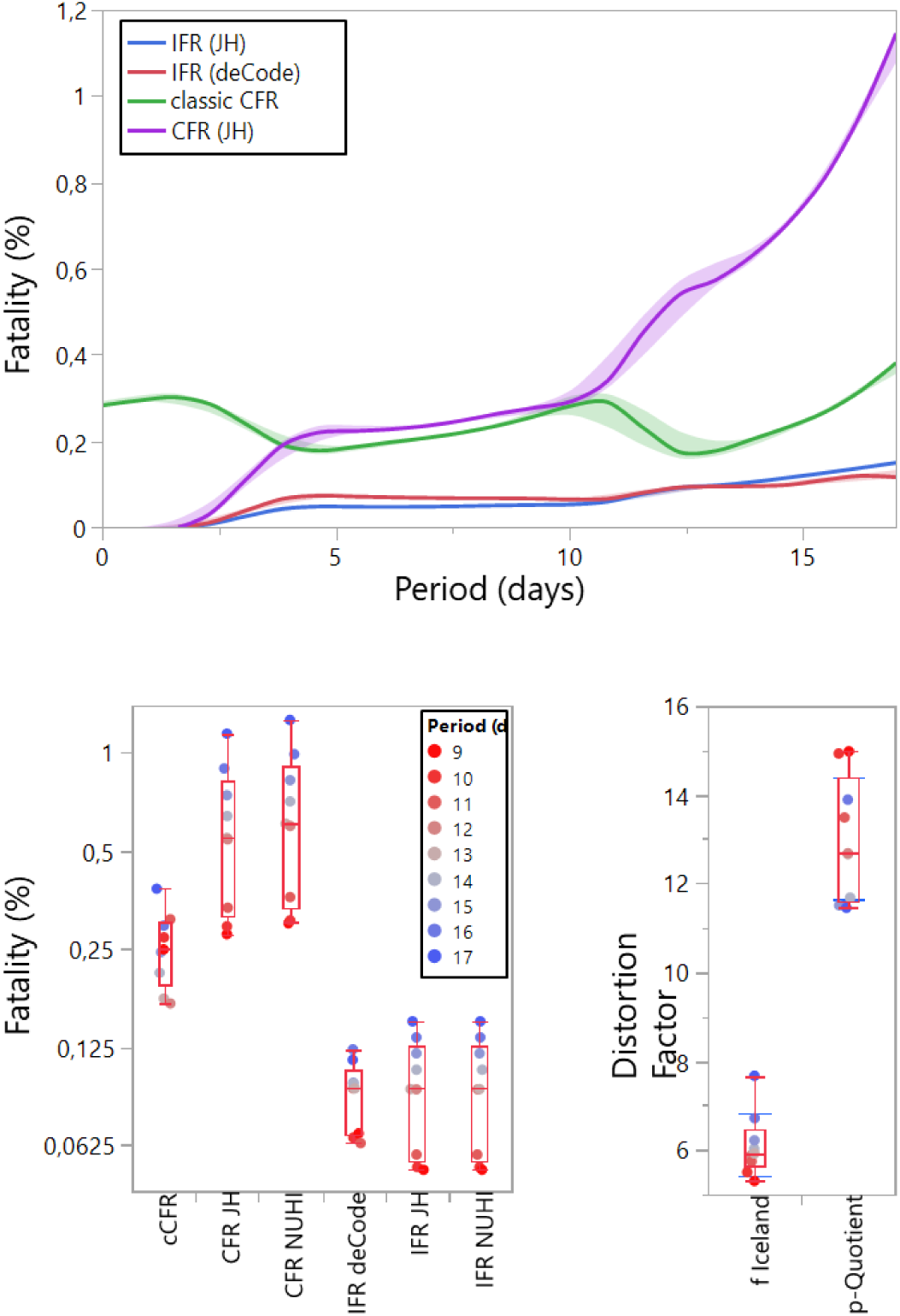
The upper panel shows four different estimates for mortality of the population of Iceland fitted by a spline function with moderate λ and 95%-CIs shaded. The IFR_deCode_ is the figure derived from testing the general population of Iceland and served to cross validate the mortality figures CFR and classic CFR that have been calculated from the data repositories of JH and the IFR that used this repository in conjunction with the test data published by Iceland’s Department of Public Health.

The lower left shows 5 different mortality figures calculated compared to the data IFR_deCode_ that is epidemiologically derived and calculated by formula (7) the data still seemed to rise with an increase in the period of the IFR as indicated by the change in colour with increasing period from red to blue. The lower left shows the comparison of the distortion factor f compared with the p-Quotient, which is the quotient for the prevalence of a positive test results in the test pool of the health officials of Iceland compared with the general population.

For the deCODE collective an epidemiologically derived prevalence of being tested positive can be calculated according to formula (2) for general population, which served to calculate an IFR_deCODE_ according to formula (7) representative for the general population and independent from the cases reported by NUHI or in the JU database. The group comparison for IFR_deCODE_ with 5 other calculated fatality rates showed a significant global group difference (n = 9, df = 5, *F* = 46.9, p = .0001), which was followed by all-pairs Tukey Kramer post hoc testing (Fig. 2). The IFR_deCODE_ with 0.088 (CI 0.067 – 0.115) did neither differ from the IFR calculated from JU data (IFR_JU_) 0.089 (CI: 0.068 – 0.117), nor the one calculated from NUHI data IFR_NUHI_ = 0.089 (CI: 0.068 – 0.117). While the classic CFR (cCFR) 0.249 (0.190 – 0.326; p = .0001), the CFR_JU_ 0.542 (CI 0.414 – 0.709, p = .0001), and the CFR_NUHI_ 0.592 (CI: 0.452 –0.774, p = .0001) tended to overestimate fatality of infection roughly 2.7-fold up to 6-fold.

This margin of overestimation is relatively low, when compared to other observed fold-differences between the CFR and the IFR in the training dataset described in Table 1.

### Composing a more comprehensive data set for testing data

In order to validate the concept of the IFR on a larger and more heterogenous collection of countries, from more continents than Europe and Asia, and in order to compare the estimates with expert consensus and conventional CFRs, a more comprehensive data set was composed. This was achieved by connecting the data from JH and ECDC up to the 31^st^ of March with the data on test numbers conducted as retrieved from the following internet sources (suppl. Table 1): data found on Wikipedia, in the COVID-19 tracking project on GitHub, the cross validated data on “Our World in Data” (OWID), and non-validated data from OWID relying on press releases for instance, but reporting its sources rigorously^18^. Double entries in different data sources were cross validated. While cross-validation indicated a high data reliability (r > .99, p = .0001), highly significantly more data that did not pass quality control of staying below a cut-off for the CFR of 10% had been retrieved from unofficial data sources (data not shown). This cannot be taken as a sign of higher inaccuracy of unofficial data per se, since the following difficulties were encountered by controlling the data on testing frequencies.

Though referenced correctly, the data in unofficial sources for one country were sometimes referring to different starting points of cumulative assessment. After finding, visiting and translating the original reports page from Japanese authorities (Supplemental Table 1) data point entries could be increased from two to 17, but it became also evident that there were data reporting cumulative test figures starting 6^th^ of March and data starting from the very beginning of case and death reporting. There is the question whether to exclude or include the cases from an international cruise ship that was under quarantine in a Japanese harbor, since most of the ship passengers were not Japanese. This is relevant for the early reporting in Japan, while, fortunately, the testing figure done on those individuals is rather negatable for later points with higher cumulative total test figures, which analyzed in the following.

Some other countries were not reporting cumulative, but solely daily, or weekly reports of their testing figures and only in their national language, which could sometimes unintendedly be misinterpreted as cumulative total, when figures were high and rapidly increasing, as for instance in Germany. Semi-official resources started to report first estimates on testing figures more than a week before official sources in Germany. These figures were too low and had been corrected by official resources, but the up to now official data are still incomplete, which can only be revealed, it one translates, reads and understands the complete report in German language (Suppemental Table 1). Moreover, reporting by unofficial sources can be sometimes more precise than official data, but points towards a new field of uncertainty. For the US, the unofficial data-tracking project on GitHub published the data differentiating in reported positive, negative and tests pending. The “test pending” category could be very relevant.

A set of 21 countries emerged (Table 3).

**Table 3:** Presented are the IFRs for the countries with their means and 95%-CIs in the validation data set. In the lines in bold, the IFRs are nested into three groups according to progressive increase in time period, ranging from below 9 days over 9-13 days to 2 weeks and more. In the two lower lines the estimates for the CFR and the classic CFR are shown.

| Country | (N) | Mean span (d) | Mean IFR (%) | CI lower | CI upper |
| --- | --- | --- | --- | --- | --- |
| Australia | 18 | 7.5 | 0.042 | 0.03 | 0.058 |
| Austria | 54 | 7.5 | 0.094 | 0.079 | 0.111 |
| Bahrain | 36 | 9 | 0.156 | 0.128 | 0.189 |
| Canada | 18 | 6 | 0.121 | 0.091 | 0.161 |
| Costa Rica | 18 | 7.5 | 0.019 | 0.012 | 0.028 |
| Croatia | 18 | 7 | 0.023 | 0.016 | 0.033 |
| Denmark | 45 | 8 | 0.14 | 0.117 | 0.167 |
| Germany | 9 | 5 | 0.067 | 0.043 | 0.101 |
| Iceland* | 54 | 7.5 | 0.036 | 0.03 | 0.043 |
| Japan | 117 | 11 | 0.054 | 0.048 | 0.061 |
| Lithuania | 9 | 6 | 0.096 | 0.063 | 0.143 |
| Malaysia | 27 | 9 | 0.052 | 0.04 | 0.067 |
| Norway | 81 | 9 | 0.059 | 0.051 | 0.068 |
| Pakistan | 9 | 8 | 0.007 | 0.002 | 0.014 |
| Peru | 9 | 6 | 0.027 | 0.015 | 0.043 |
| Poland | 36 | 6.5 | 0.086 | 0.07 | 0.105 |
| Russia | 18 | 7.5 | 0.039 | 0.028 | 0.053 |
| Slovenia | 45 | 10 | 0.148 | 0.124 | 0.176 |
| South Korea | 162 | 13.5 | 0.06 | 0.055 | 0.067 |
| Switzerland | 9 | 6 | 0.158 | 0.106 | 0.232 |
| Thailand | 27 | 8 | 0.021 | 0.015 | 0.028 |
| **IFR<9d** | **374** | **< 9 days** | **0.041** | **0,037** | **0.044** |
| **IFR 9-13d** | **299** | **9-13 days** | **0.087** | **0.080** | **0.095** |
| **IFR> 13d** | **146** | **≥ 2 weeks** | **0.097** | **0.086** | **0.109** |
| CFR | 91 | 12 | 2.73 | 2.28 | 3.26 |
| classic CFR | 91 | - | 0.680 | 0.568 | 0.814 |

### Comparison of the IFRs with the CFRs of 21 countries with minimal essential information on testing

By data mining, I was able to retrieve the following data on cumulative test numbers. 129 data points came from the national official reporting organ of the countries, 14 data points were originally retrieved via OWID, but controlled, and then updated with official national data, 34 data points came from OWID, 19 for the USA from the tracking project, where I relied on the confirmed test numbers, excluding the pending ones, to avoid partial doubling of data. Finally, 104 data points were retrieved from Wikipedia on the COVID-19 pandemic information pages, where also pdfs and links to the national sources of data are published. These data points belonged originally to 53 countries, but only 21 fulfilled inclusion criteria.

These countries are listed with their IFRs in Table 3, which also provides means and 95%-CIs for the means of Japan and Korea are both at the end of a consolidation phase and did neither show values for the classic CFR nor for CFR in line with expert estimates, but their IFR estimates are again in line with that of all other countries (Fig. 3). The IFR is with 0.05-0.3 a bit lower than the current expert consensus, but the margin reflected (Fig.3 and Table 2) is the narrowest for all countries.

**Fig.3:**
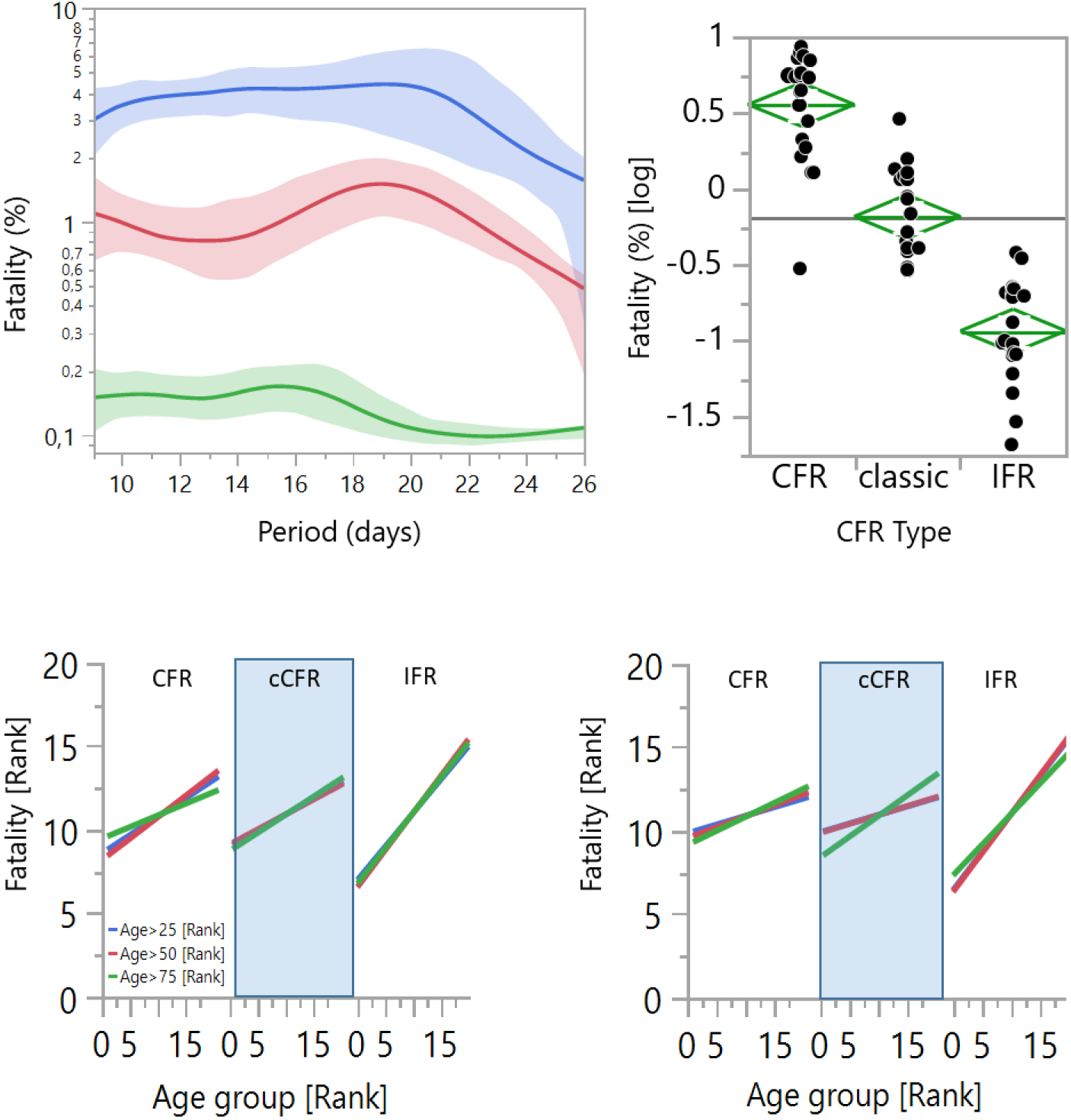
In the upper left the CFR (blue) classic CFR (red) and IFR (green) are compared in relation to the length of the period between reported deaths and cases. Spline of the Means line and 95%-CIs shaded are shown. The upper right compares the means and diamonds with their peaks representing 95%-CIs for the log-normalized data of the 21 countries with the CFRs and IFRs from periods >9 days. The lower panels show on the left hand side comparisons of the ranks of the proportions for three different age groups (above 25, 50 and 75 years) in males and on the right hand side in females.

The median for all 21 countries for the IFRs was 0.11 (95%-CI: 0.073–0.15) and significantly different from CFR with 3.59 (95%-CI: 2.57–5.00, p = .0001) and the classic CFR of 0.66 (95%-CI: 0.47–0.91, p = .0.0001; Figure 3 upper left).

An epidemiologically derived IFR of 0.040 % (95%-CI: 0.029%–0.055%) was determined for Iceland and was very close to the calculated IFR of 0.057% (95%-CI: 0.042–0.078), but highly significantly 2.7–6-fold lower than CFRs. IFRs, but not CFRs, were positively associated with the medians of the countries IFRs were significantly positively associated in men with the proportion of elderly people in the respective countries age cohorts >25 years (r = 0.44, p = .044), >50 years (r = 0.47, p = .034), > 75 years (r = 0.46, p = .036), while only significantly associated with the age cohort >75 years (r = 0.45, p = .041) in females (Figure 3 lower part).

## Discussion

During the outbreak of a pandemic, it is difficult to estimate and then communicate a CFR precisely enough from epidemiological data^5^, while situations, where many countries may run out of health resources nevertheless require guidance and recommendations by experts^1,11^. In the current situation of the SARS-CoV-2 outbreak, the general public is confronted with large differences in the estimation of conventional CFRs between countries like Italy (>7%), South Korea (>1%) or Germany (0.4%). Experts’ estimates for CFR vary between 0.1 up to 1, which reflects a broad range of pandemic scenarios and a broad range of possible mitigation and suppression strategies which could be derived by experts. In this situation, modelling of scenarios is applied, by relying on key parameters of epidemic spread^1,9,11^. The most crucial imputation for such models is the basic reproductive number R_0,_ which can be assessed from Data on the early outbreak of a pandemic, but is compromised by a significant level of uncertainty on top of any epidemiologically derived level of statistical confidence due to essential uncertainties how such data can be transferred from one scenario to another. To put this in simple terms, a cruise ships’ field conditions during a quarantine^8^ are not comparable to an unanticipated new pandemic outbreak in China^9^, are not comparable to Europe^1^. Applying modelling now requires even more imputation on the latent period of the infectious period and the interval of half-maximum infectiousness. Values could again all be derived from environmental observations, and all prone to a substantially unknown extend of error, especially if assessed for a new pathogen for the very first time. In principal, imputation values can be subject to modelling itself^21^. Also modeled values used for imputation into the next model as an assumption will not avoid further inflating the level of uncertainty^12^. What will come out at the end, is the most precise we can get with modeling, we will end up in a range of scenarios from the Spanish Influenza down to the seasonable flu and with a range of mitigation or suppression strategies that will all be supportable, principally. To improve modelling substantially would now require narrowing the range of fatality down to a margin at which modelling makes sense. A very recent publication describes a way, how we could achieve this goal^14^. In this publication, again a high number of imputations had to be fed again into a model, again field conditions, which are not comparable between each other, had to be chosen and basic assumptions had to be made to model an estimate for the IFR. While this estimate may indeed be more precise than former estimates, the basic problem of requiring a lot of proper field work and requiring a lot of as precise basic assumptions as possible to avoid excessive error propagation of unknown extend, had not been solved or dealt with. The IFR was adjusted for census and for a problem, which at first glance my IFR is not capable to cope with, ascertainment aspects^14^. However, a more morbid person, which ends up preferentially in a test pool during the outbreak of a pandemic, may not be a more elderly person or a person having better access to testing, only. We just are not aware, of the many factors that all may contribute to preferentially testing certain people in the heat of a pandemic. We are trusting in *ex posteriori* - derived assumptions and confirm them with a model.

Here I propose a way to adjust for this one particular problem in infection epidemiology – preferential selection of persons that will show up in a test pool -, if there was equal access to the test pool and enough testing capacity.

I deduced that my approach only required sequentially monitored confirmed cases, recovered cases, and death events in conjunction with total numbers of diagnostic tests performed in a given population. These data, except the total number of tests conducted, are already subject to official reporting and data collection by national and international centres of disease control. I showed that this approach successfully stabilized against selection bias, with a validation against field data in Iceland and by comparing CFRs and calculated IFRs for plausibility and for their ability to reflect an association with census not only within countries, but also across countries. The latter is important, if we assume that biological aspects of a virus are valid across the boundaries of nations.

This approach required deducing a correction for selection bias, here termed *f*, and validating the effect of applying this correction factor to empirical data, to arrive at preliminary estimates for 21 countries of the world with 95% confidence intervals for IFRs. It is a preliminary estimate with all its shortcomings, but it is a single, potentially relevant variable for crude mortality, logically deduced, requiring only data imputations that potentially could be delivered with high certainty during future pandemics, with low effort, and at low cost. More crucially, it does not require exponential modelling or substantial expert knowledge to arrive at a readout for crude mortality that appears to be robust between countries and appears to reflect viral biology.

The correction factor *f* is simply the square root of the quotient of the total population divided by total tests conducted. Even if countries would either go through periods of rapid test rate growths or experience limitations with their testing capacities, the distortion provoked will not lead to huge uncertainty ranges by a substantial unknown error propagation.

Correcting CFRs with *f* is capable of harmonizing differences in CFRs between countries that would otherwise be difficult to explain. Amongst these candidate countries are Japan, South Korea, Iceland, and Norway, which have done meticulous work in dealing with their testing data, protocolling everything transparently and timely, to the public, and moreover, which have strong economies and strong health care systems to cope with the current pandemic.

Amongst those, that report their testing data almost in real time and comprehensively, is Pakistan. Pakistan is a country, which seems to fall out of the range of IFRs, with an IFR of 0.007 that is roughly 10-fold lower than the one reported for the so-called developed countries. Since testing was reported transparent and timely, it is important to understand, whether this extremely low IFR figure reported in Table 3 could be possibly realistic, or not.

Population statistics of this country compared to any of the developed countries is very informative with this regard. As of 2018, 6.7% of the population in Pakistan were over 60 years old and 45% were younger than 20. In Germany 19 % were younger than then 20 years old, 29% were older than 60 years. The CFR for people aged 60-69 compared to people aged below 20 has been published to be roughly 20-fold higher^9^. Even though this figure is most likely too high because of the CFR being prone to be inflated by selecting a more morbid population into the testing pool, there is agreement amongst scientists, that SARS-CoV-2 at least shows a strong difference within countries or within regions to be associated with higher values for older people^4^. By using the correction factor *f* it is now possible to show significant association of a SARS-CoV-2 related mortality figure with age composition not only within, but also between countries.

A crucial problem for testing data prevails, on the present level of accuracy for official reporting. Even for countries like the US, Italy and the UK, with very timely and detailed test reporting, a calculation of an IFR could not have delivered any meaningful outcome than a CFR calculation, or essentially, guessing. In the heat of an ongoing pandemic, it is often still possible to report a death almost in real-time. At the same time, cases will be prone to unreported delay factors, once testing reaches maximum testing capacity. During an exponential growth, this will cause a severe distortion, if a CFR or IFR is calculated. This might happen just because we think that our cases are reported with the day of testing, but in fact, the case may appear as a reported case many days after the death of the person, leading to a CFR of sometimes more than 1,000% (for an example; UK, Figure 1).

On the opposite, Germany had been able to expand its testing volume presumably (pending data revision as of April 6^th^, suppl. Table 1 for reference) by a factor of 2.7 from week 11 to week 12 of the current year, which inflated case number.

Such bias will not only limit the validity of the CFR, but also limit the validity of IFR calculation. However, in contrast to the problems of unknown error propagation in modeling approaches, such limitation could principally be dealt with. if a similar pandemic outbreak, with concomitantly high enough testing capacity, in a well enough informed and health educated cohort of enough people would occur again. The numbers generated here for the IFRs need to be critically taken into consideration by abductive reasoning in an interdisciplinary committee of experts. They are no standalone figures for mortality, since they mainly reduce one particular sort of bias amongst the manifold in empirical work in the field of infection epidemiology. With the decisions to follow certain mitigation or suppression strategies by almost all developed nations, there will be problems to be solved around the globe^22^.

There is one last question, which I will not discuss here. Provided that there was a defined place with more than 364.000 people, and provided at that place everybody knew what COVID-19 is, will not miss a single death, will take care of people coughing, and provided that there was enough testing capacity at hand: Could it be that my formula (5) to correct for selecting more morbid persons into a test pool, will also correct for all other sorts of selection bias, therefore delivering an ultra-precise early estimate for mortality?

## Data Availability

All data used for this study are freely available and accessible via the given references. Composed datasets are available upon request

## Competing Interests

None to be declared.

## Funding Statement

No funding supported the work presented.

## Acknowledgement

I would like to thank the Health Authorities of Iceland and deCODE genetics for making their data freely available for the public. Many thanks to the Open Data Source Community in its broadest sense. There is massive amounts of work going into curation of pages like the one we can now find on Wikipedia about testing. I thank my colleagues, dear friends, and family members, who critically read the manuscript.

## Notes

### Competing Interest Statement

The authors have declared no competing interest.

